# A Research on the Results of Viral Nucleic Acid Tests and CT Imaging Variation of Patients with COVID-19

**DOI:** 10.1101/2020.05.08.20037556

**Authors:** Meng Xu, Xun Liu, Chuhong Su, Yuping Zeng, Jinqian Zhang, Xuwen Li, Guirong Liu, Jinjun Xie, Hongyong Liu, Yusheng Jie

**Affiliations:** Department of Infectious Diseases the Third Affiliated Hospital of Sun Yat-sen University Guangzhou P.R. China; Department of Hospital Infection Management, the Third Affiliated Hospital of Sun Yat-sen University Yuedong Hospital, Meizhou, P. R. China; Clinial data center, the Third Affiliated Hospital of Sun Yat-sen University, Guangzhou, P. R. China; Department of Nutrition and Food Hygiene, Southern Medical University, Guangzhou, P. R. China; Department of Intensive Medical, the Third Affiliated Hospital of Sun Yat-sen University Yuedong Hospital, Meizhou, P. R. China; Department of Cardiovascular Medicine, the Third Affiliated Hospital of Sun Yat-sen University Yuedong Hospital, Meizhou, P. R. China; Department of Laboratory Medicine, The Third Affiliated Hospital of Sun Yat-sen University Yuedong Hospital, Meizhou, P. R. China; Department of Radiology, The Third Affiliated Hospital of Sun Yat-sen University Yuedong Hospital, Meizhou, P. R. China; Department of Renal Medicine, the Third Affiliated Hospital of Sun Yat-sen University Yuedong Hospital, Meizhou, P. R. China

**Keywords:** COVID-19, Diagnosis, Virus Nucleic Acid, Lung CT

## Abstract

**Background:** Coronavirus disease 2019 (COVID-19) has become a global health problem. We aim to investigate the changes in the results of viral nucleic acid tests on pharyngeal swabs and feces of patients with COVID-19 and CT imaging of lungs as the disease progresses.

**Methods:** Seven patients with COVID-19 in the third affiliated hospital of Sun Yat-sen University Yuedong Hospital were retrospectively enrolled with clinical features, including imaging staging, and performance characteristics of viral nucleic acid test results of pharyngeal swabs and feces. The dynamic changes of these features were observed during hospitalization, and therapeutic effect and prognosis of patients were evaluated.

**Results:** The results of seven cases with COVID-19 were positive for viral nucleic acid tests on pharyngeal swabs early after the onset of symptoms, and then turned negative; while the results of viral nucleic acid tests on feces were persistently positive in the mid-term clinical treatment and recovery period. And the viral nucleic acid test results were capricious in three cases. Pulmonary CT imaging showed characteristic changes in early, advanced and recovery phases.

**Conclusion:** The application of viral nucleic acid detection and pulmonary CT imaging can be used for screening of suspected cases. Fecal nucleic acid test should be recommended as the reference of discharge standard, in order to minimize the risk of transmission from digestive tract.

## 1. Introduction

Beginning in December 2019, a cluster of unidentified respiratory illness was successively discovered in Wuhan, China, and subsequently spread in various provinces of China and over 90 countries worldwide. The World Health Organization (WHO) named this kind of unidentified disease as COVID-19. Recent researches show that severe acute respiratory syndrome coronavirus 2 (SARS-CoV-2), a β-type coronavirus, has more than 85% homology with bat SARS-like coronavirus, acting as the causative organism of COVID-19, and it is easily transmissible in humans. The source of infection is mainly infected patients, and also asymptomatic patients. Respiratory droplets and close contact transmission are definite transmission routes and there is a possibility of aerosol transmission in a relatively closed environment when exposed to high concentrations of aerosol for a long time. Primary manifestations of COVID-19 include fever, dry cough, limb weakness and dyspnea[1]. The latest research found that COVID-19 cases may also show gastrointestinal symptoms, but the incidence is low[2]. Similarly, clinical manifestation of cases in our hospital also include gastrointestinal symptoms. By March 5, 2020, on the verge of reaching 100000 confirmed cases of COVID-19 had been reported in China and other countries around the world [3], and 1524 patients had died, equivalent to a mortality rate of about 2%[4].

In addition to the corresponding epidemiological history and clinical manifestations, confirmed cases of COVID-19 need to have etiological evidence, which means positive real-time reverse-transcription-polymerase-chain-reaction (RT-PCR) results for the detection of SARS-CoV-2 nucleic acids or highly homologous viral gene sequencing to known SARS-CoV-2[5]. Compared with viral gene sequencing, viral nucleic acid detection is more cost-effective, efficient, and faster, making it the main method for pathogenic diagnosis. Due to the lack of nucleic acid detection kit and its long detection time span characteristics, viral nucleic acid detection for large number of patients in the early stages of outbreak becomes difficult. Some clinicians proposed that the pulmonary computerized tomography (CT) scan of the lungs should be included in the diagnostic criteria, and even discussed whether it can be used as a diagnostic basis for confirmed cases[6].

However, the insistency of viral nucleic acid tests and CT scan results triggered controversy. This article analyzes the epidemic history, nucleic acid testing results, and CT images of 7 confirmed cases admitted to our hospital, and discusses the changes in nucleic acid tests and CT images as the disease proceeds, in order to provide scientific basis for screening, early diagnosis and dynamic assessment of the disease.

## 2. Methods

### 2.1 Study design and participants

This was a retrospective analysis study of the clinical features of 7 patients with COVID-19 who were confirmed COVID-19 pneumonia in the third affiliated hospital of Sun Yat-sen university Yuedong hospital, from January 2020 to February 2020. Covid-19 patients were diagnosed and defined the degree of severity on the basis of the Diagnosis and Treatment of COVID-19 pneumonia (Trial Edition 7). This study was approved by the Ethics Committee of the third affiliated hospital of Sun Yat-sen university Yuedong hospital [2020–01]. Written informed consent was waived owing to the rapid emergence of this infectious disease.

### 2.2 Virus nucleic acid detection

Virus nucleic acid detection was performed by RT-PCR (BIOER, China and Shanghai ZJ Bio-Tech, China). The samples were pharyngeal swabs and fecal specimens of patients, detecting the RNA-dependent RNA polymerase, nucleocapsid protein (N) gene and envelope protein (E) gene of SARS-CoV-2. The test results were classified as negative and positive.

### 2.3 CT scan

All CT images were obtained with patients in the supine position and scanned from the level of the upper thoracic inlet to the inferior level of the intercostal angle using a Brilliance 16-slice CT (Philips, The Netherlands). The imaging results were interpreted and reviewed by 2 attending or higher-level radiologists.

## 3. Results

### 3.1 Epidemiology

The patients were 7 newly confirmed cases of COVID-19 pneumonia by the pharyngeal swab nucleic acid test from January 25 to February 6, 2020 in the Third Affiliated Hospital of Sun Yat-sen University Yuedong Hospital, and 5 were discharged from hospital on February 23, 2020, and the other 2 cases are still hospitalized. All cases were named as case plus number for convenience, such as case 1. Among them, Cases 1, 3, 4, 5 and 7 are returnees from Wuhan city, Hubei Province. Cases 2 and 6 are close contacts of confirmed cases. Case 2 is a colleague of a confirmed patient from other hospital, and Case 6 is the mother of Case 4. The demographics of patients is shown in Table 1.

**Table 1.** Demographics of confirmed cases of COVID-19

| **Basic characteristics** | **N ( % )** |
| --- | --- |
| **Age** |  |
| 0 ~ | 0 ( 0 ) |
| 20 ~ | 3 ( 42.9% ) |
| 40 ~ | 3 ( 42.9% ) |
| 60 ~ | 1 ( 14.2% ) |
| ≥80 | 0 ( 0 ) |
| **Sex** |  |
| Male | 3 ( 42.9% ) |
| Female | 4 ( 57.1% ) |
| **Habitual residence** |  |
| Hubei Province | 5 ( 71.4% ) |
| Guangdong Province | 2 ( 28.6% ) |
| **Contact with people from**<br>**Wuhan** |  |
| Positive | 4 ( 42.9% ) |
| Negative | 3 ( 57.1% ) |

### 3.2 Clinical Manifestation

All patients had respiratory symptoms, mainly cough and pharynx discomfort, throughout the course of the disease, accompanied by fever in 3 patients and limb weakness in 6 patients. Abdominal pain, diarrhea and other gastrointestinal symptoms were also observed in 3 patients. The clinical symptoms of all cases are shown in Table 2.

**Table 2.** Clinical features of patients with COVID-19

| **Clinical features** | **N ( % )** |
| --- | --- |
| **Type of case** |  |
| Mild | 0 ( 0 ) |
| Common | 6 ( 85.7% ) |
| Severe | 1 ( 14.3% ) |
| Critical | 0 ( 0 ) |
| **Fever** |  |
| Positive | 3 ( 42.9% ) |
| Negative | 4 ( 57.1% ) |
| **Nasal Obstruction** |  |
| Positive | 4 ( 57.1% ) |
| Negative | 3 ( 42.9% ) |
| **Catarrh** |  |
| Positive | 1 ( 14.3% ) |
| Negative | 6 ( 85.7% ) |
| **Rhinobyon** |  |
| Positive | 2 ( 28.6% ) |
| Negative | 5 ( 71.4% ) |
| **Productive Cough** |  |
| Positive | 2 ( 28.6% ) |
| Negative | 5 ( 71.4% ) |
| **Dry Cough** |  |
| Positive | 5 ( 71.4% ) |
| Negative | 2 ( 28.6% ) |
| **Chest distress** |  |
| Positive | 3 ( 42.9% ) |
| Negative | 4 ( 57.1% ) |
| **Polypnea** |  |
| Positive | 2 ( 28.6% ) |
| Negative | 5 ( 71.4% ) |
| **Limb weakness** |  |
| Positive | 6 ( 85.7% ) |
| Negative | 1 ( 14.3% ) |
| **Diarrhea** |  |
| Positive | 3 ( 42.9% ) |
| Negative | 4 ( 57.1% ) |
| **Stomachache** |  |
| Positive | 2 ( 28.6% ) |
| Negative | 5 ( 71.4% ) |

### 3.3 The results of virus nucleic acid detection

Virus nucleic acid test results are shown in Table 3. All cases were viral nucleic acid-positive in pharyngeal swabs after admission, and subsequently turned negative. However, Cases 1, 2 and 6 were positive again for viral nucleic acid tests on Day 12, 9 and 12 after the onset of symptoms, respectively, of which Case 6 was positive again after being negative for 3 consecutive times. Moreover, Case 5 remained positive for the pharyngeal swab virus nucleic acid test during the first 20 days after the onset of symptoms. Besides, we got positive results in faeces of 4 patients (Cases 1, 3, 4 and 6) while no positive results were detected in faeces of the other 3 patients (Cases 2, 5 and 7). The viral nucleic acid test by faeces of Case 6 was positive again after 4 consecutive negatives. Case 4 remained positive for faeces virus nucleic acid test during the first 22 days after the onset of symptoms.

**Table 3.**
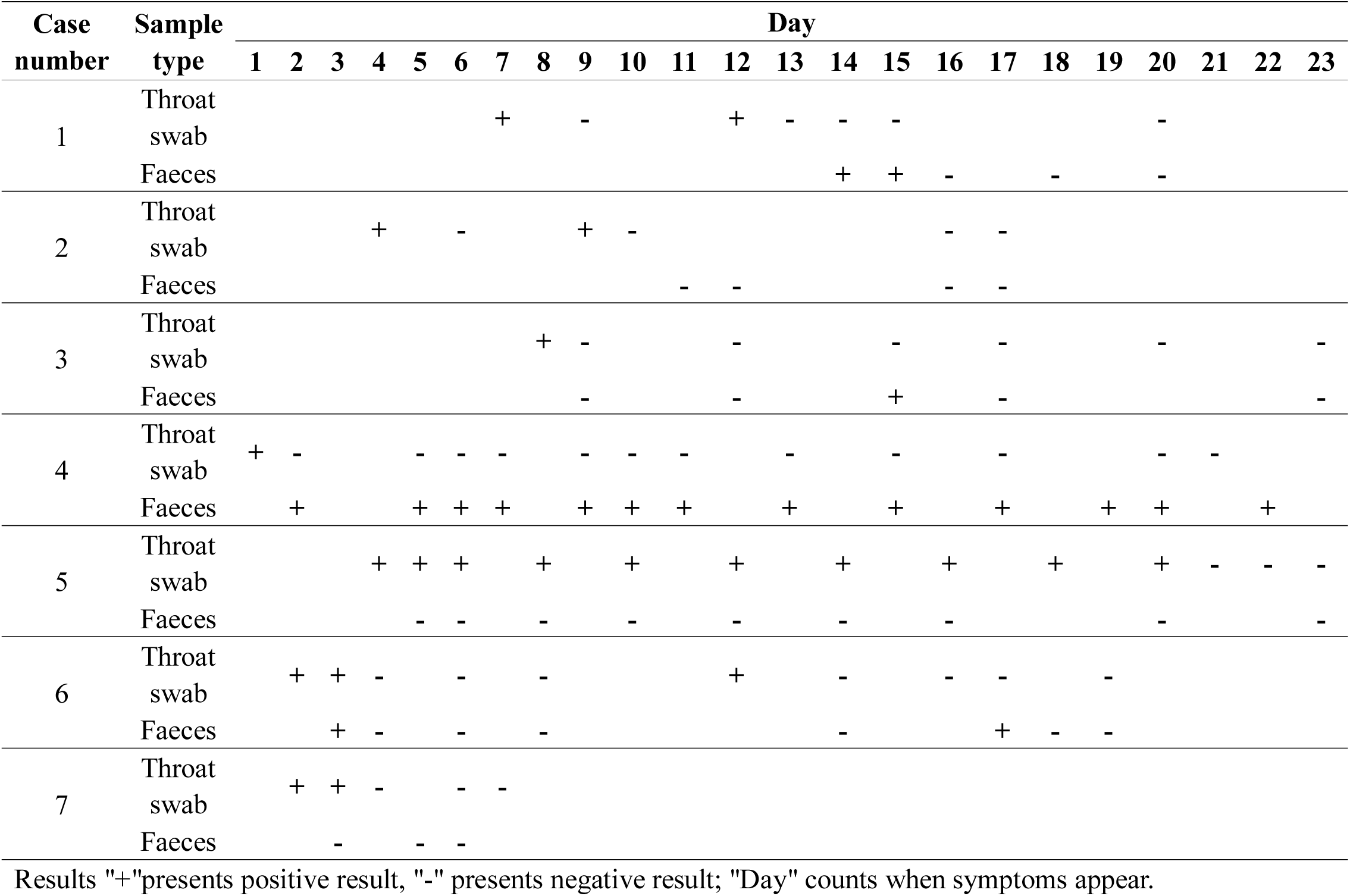
Viral nucleic acid test results of COVID-19 cases

### 3.4 The results of CT examination

Lung CT images revealed the evidence of pneumonia in all 7 cases (Table 4 and Figure 1). Except for Case 7, whose lower lobe of right lung was only involved. all patients had bilateral lung involvement. Five cases (Cases 1,2,3,4,6) showed peripheral distribution, while the other 2 cases showed diffused pulmonary abnormality. The majority of patients had characteristic ground-glass opacity (GGO) in CT images of the lungs and a few patients showed thickened blood vessel (case 6) and interstitial tissue (case 5). No pulmonary consolidation or cystic changes was observed.

**Table 4.** Imaging changes of CT examination of COVID-19 cases

| Case Number | Location of Lesions | Type of Lesions |  |  |  |
| --- | --- | --- | --- | --- | --- |
|  |  | GGO | Consolidation | Blood Vessel Thickening | Interstitial Tissue Thickening |
| 1 | Lower lobes of both lungs | - | - | - | - |
| 2 | Upper lobe of left lung;<br>Lower lobes of both lungs | + | - | - | - |
| 3 | Lower lobe of both lungs | + | - | - | - |
| 4 | Upper lobe of right lung;<br>Lower lobes of both lungs | - | - | - | - |
| 5 | Upper lobe of left lung;<br>Lower lobes of right lungs | + | - | - | + |
| 6 | Lower lobe of both lungs | + | - | + | - |
| 7 | Lower lobe of right lungs | + | - | - | - |

**Figure 1.**
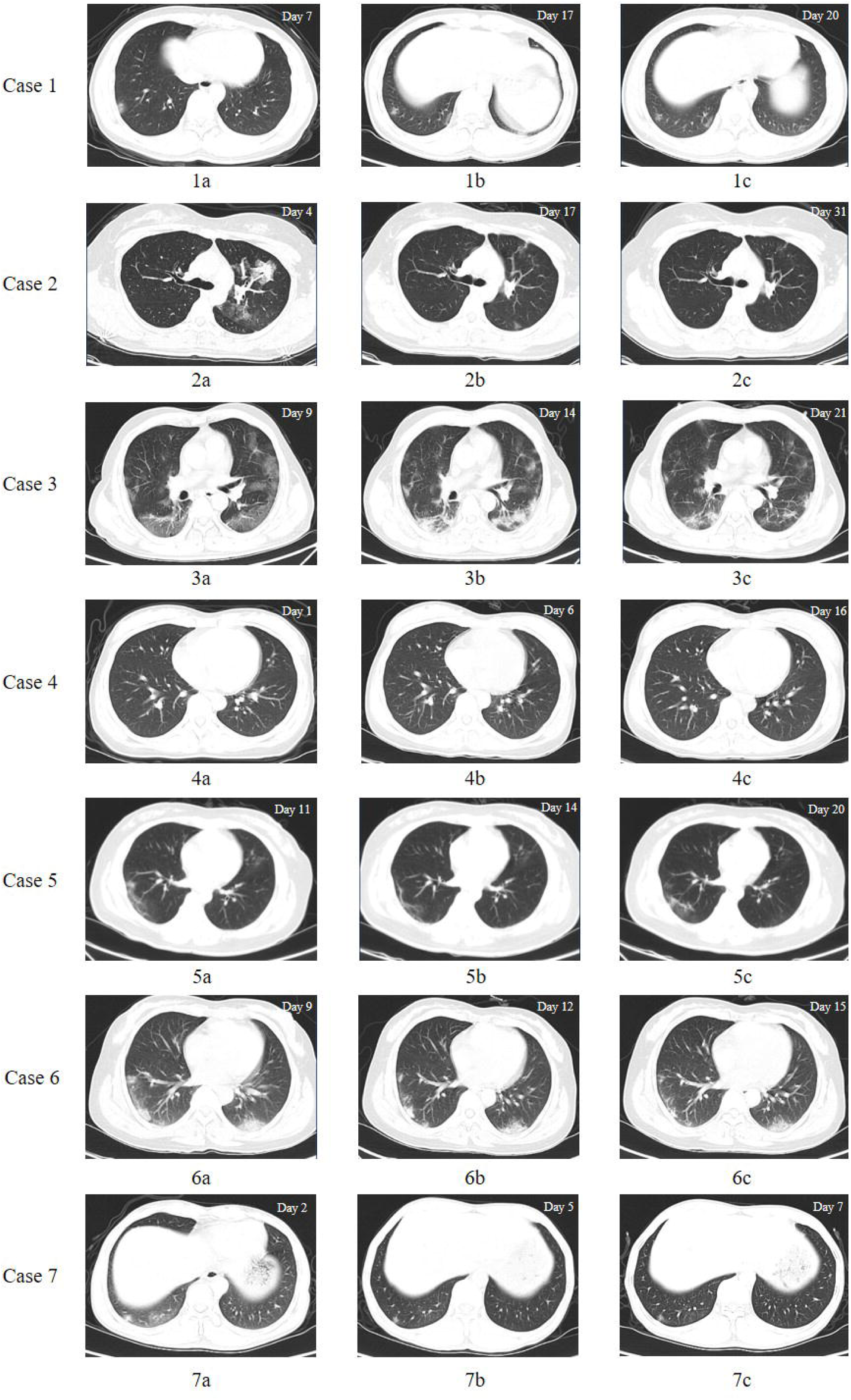
Pulmonary CT images of confirmed cases of COVID-19. The pulmonary CT scan was conducted from the upper level of thoracic inlet to the inferior level of the costophrenic angle with all 7 confirmed patients in the supine position. Representative pictures were presented out of all pictures from every pulmonary cross-section in each patient.

On the 7^th^ day, a chest CT examination of Case 1 was conducted in our hospital, which showed obvious lung marking changes, and multiple patchy shadows under the pleura in the lower lobe of the lungs (Figure 1-1a). The result got worse on day 10 (Figure 1-1b) until it became better again on day 20(Figure 1-1c). CT image showed that Case 2had multiple patchy shadows with increased density and fuzzy margin scattered in the left lung on day 4, which were mainly distributed in superior lobe of left lung(Figure 1–2a to 2c), and later slowly disappeared. CT images of Case 3 and Case 4 (Figure 1–3a to 4c) were similar, which started with multiple patchy slightly high-density shadows, and later absorbed on day 14 and day 6, respectively. In addition, we found that Case 5 (Figure 1–5a to 5c) demonstrated fluctuating lung CT images, which showed earlier alleviated and later aggravated manifestation. Case 6 (Figure 1–6a to 6c) showed GGO with fuzzy margin in the lower lobe of both lungs, with focus of infection near the pleura and thicken blood vessels nearby. Case 7 (Figure 1–7a to 7c) showed a slight increase in the markings of both lungs, and GGO were seen in the outer basal and posterior basal segments of the lower lobe of right lung.

## 4. Discussion

The current pathogenic detection methods of COVID-19 mainly include fluorescent RT-PCR detection of virus nucleic acid or virus gene sequencing. The specimens that can be collected include pharyngeal secretions, sputum, bronchial veolar lavage fluid, blood, faeces, urine, etc. Test methods, specimens, test reagents, and test operations are all factors that may affect the results. Our hospital uses fluorescent RT-PCR to detect virus nucleic acid. The specimens are mainly pharyngeal swab and faeces. For the seven confirmed cases in our hospital, virus nucleic acid test results mainly showed that the pharyngeal swab virus nucleic acid test results were positive in early stage after the onset of symptoms, and then turned negative while the faeces test results were persistently positive in the middle of clinical treatment and recovery stage. Among them, the pharyngeal swab test result of case 6 returned positive again after being negative for 3 consecutive times, and faeces specimens test result returned to positive for being negative for 4 consecutive times, which indicates that the test result may be related to the amount of virus in the body, the sensitivity of the test kit, and the repeated illness of COVID-19. This situation was consistent with what was reported in the press conference of the government in Guangdong Province where 14% of discharged patients were "Recurrence". The faeces test results of Case 4 and the pharyngeal swab test results of Case 5 remained positive for virus nucleic acid test even on day 20.Studies have found that there is a strong binding affinity between the S protein of SARS-CoV-2 and the human angiotensin receptor 2 (ACE2)[7]. ACE2 is also highly expressed in the absorptive epithelial cells of the colon and ileum, indicating that the digestive system is a potential route of infection[8],and the possibility of gastrointestinal transmission in case 4 is not ruled out. Moreover, Chinese scientists have isolated the virus from the feces, the damaged mucous membranes and bleeding of the gastrointestinal tract, suggesting that more attention should be paid to preventing fecal-oral transmission[2].

The visual findings from COVID-19 cadaver studies are consistent with the distribution of imaging changes, considering that the GGO images seen in imaging are corresponding to the lung lesions seen in visual findings [9]. CT scan has the advantage of high image resolution, and the early lung CT manifestations of COVID-19 have certain characteristics, which plays a very important role in early diagnosis and treatment of COVID-19[10]. According to the scope and type of the lesion, the lung CT manifestation is divided as early, advanced, severe and critical, and recovery stages[11]. Confirmed cases in our hospital include image changes in the early, advanced and recovery phases. Until the end of February 23, CT changes were seen in both lungs of 6 cases among the 7 confirmed cases, the basal segment of the lower lung lobe in 5 cases was involved, 5 cases showed GGO, and 1 case had thickened blood vessels and thickened interstitial tissue. No pulmonary consolidation occurred in these 7 cases.

Combined results of nucleic acid test and CT imaging among the 7 confirmed patients indicate that the 2 detection methods are comparable. Both detections were positive in 6 cases. Nucleic acid tests were negative and CT changes were positive in 1 case, and nucleic acids were positive after re-examination. No case with both negative by the 2 methods was observed, and the case with positive nucleic acids and negative CT changes was not found. In general, the positive results of the pharyngeal swab tests came earlier than the CT image indicating pneumonia alleviation. Studies have found that CT imaging changes in the lungs are more sensitive to the detection of COVID-19 than pharyngeal swab viral nucleic acid tests[12].

Through comparison, it was found that virus nucleic acid test and CT examination each have their own sensitivity. Based on the gold standard for viral pathogen detection, CT imaging changes in the lungs are carried out to a certain extent in the prevention and control of suspected patients and the monitoring of disease progress. As one of the designated hospitals in Guangdong Province and one of the key hospitals for fever diagnosis, our hospital performs viral nucleic acid detection and lung CT at the same time during the prevention and control of the coronavirus outbreak. In the early phase, faeces viral nucleic acid tests were also conducted besides pharyngeal swab tests for viral nucleic acid detection, actively investigating suspicious patients in our hospital from various aspects to reduce the risk of nosocomial infections.

## 5. Conclusion

In the diagnosis of COVID-19, viral nucleic acid test is used as the pathogenic evidence. With unknown nucleic acid test result and obvious symptoms and signs, the lung CT examination can be used as assistance to diagnosis so as to reduce missed diagnosis and misdiagnosis. In confirmed patients, CT changes in the lungs can assist in disease assessment. For patient discharge, fecal nucleic acid test can be used to prevent the patients with negative throat swab viral nucleic acid and positive fecal nucleic acid from recrudescence or becoming potential infectious sources.

## Data Availability

no data available.

## Availability of data and materials

The datasets used and/or analyzed during the current study are available from the corresponding author on reasonable request.

## Ethical approval

This study was approved by the Ethics Committee of the third affiliated hospital of Sun Yat-sen university Yuedong hospital [2020–01]. Written informed consent was waived owing to the rapid emergence of this infectious disease.

## Abbreviations

COVID-19: oronavirus disease 2019
WHO: The World Health Organization
SARS-CoV-2: severe acute respiratory syndrome coronavirus 2
RT-PCR: real-time reverse transcription-polymerase chain reaction
CT: computerized tomography
GGO: ground-glass opacity
ACE2: angiotensin-converting enzyme 2

